# Generation of novel SARS-CoV-2 variants on B.1.1.7 lineage in three patients with advanced HIV disease

**DOI:** 10.1101/2022.01.14.21267836

**Authors:** Anna C. Riddell, Beatrix Kele, Kathryn Harris, Jon Bible, Maurice Murphy, Subathira Dakshina, Nathaniel Storey, Dola Owoyemi, Corinna Pade, Joseph M. Gibbons, David Harrington, Eliza Alexander, Áine McKnight, Teresa Cutino-Moguel

**Author notes:** **Correspondence:** Anna Riddell and Teresa Cutino-Moguel.

## Abstract

The emergence of new SARS-COV-2 variants is of public health concern in case of vaccine escape. Described are three patients with advanced HIV-1 and chronic SARS-CoV-2 infection in whom there is evidence of selection and persistence of novel mutations which are associated with increased transmissibility and immune escape.

## BRIEF REPORT

Chronic SARS-CoV-2 infection persisting for many months has been described in numerous immunocompromised patients since January 2020 (1, 2). In these patients with prolonged and partially or uncontrolled virus replication (due to their immune deficit), there is an optimal environment for virus evolution and the generation of novel variants (3). This is a concern for the pandemic global response due to the possibility of vaccine escape: where novel mutations in the virus genome (in particular the spike protein (S)) lead to reduced susceptibility to nAb. Novel variants with accumulated mutations in the S gene (alpha, beta and delta) have already been shown to have reduced susceptibility to neutralising antibodies (nAb) in relation to wild type (WT) Wuhan 1-Hu-1 (4, 5). nAb are predictive of immune protection from SARS-CoV-2 and correlate with virus clearance and survival (6).

Here we present three patients with advanced HIV-1 (CD4 cells less than 200 × 10^6^/L) and chronic SARS-CoV-2 infection in whom we have detected the emergence of novel mutations *in vivo* via Next Generation Sequencing (NGS). In one patient, we also describe the neutralising antibody response.

Patient A, in their 40s, was admitted in early 2021 with shortness of breath, significant weight loss, fevers and night sweats. SARS-CoV-2 infection was detected via a RT-PCR of a combined nose and throat swab (CNTS). A new diagnosis of HIV-1 was confirmed during the admission. Chest radiograph changes were consistent with miliary *Mycobacterium tuberculosis* rather than COVID-19 pneumonia. *Pneumocystis jirovecii* (PCP) prophylaxis and empirical anti-tuberculous treatment with Rifater®, pyrazinamide, ethambutol was commenced. Antiretroviral therapy (ART) was started after discharge. During a subsequent admission over two months later, a CNTS was negative for SARS-CoV-2 RNA by RT-PCR.

Patient B is also in their 40s and has a history of diffuse large B cell lymphoma and HIV-1. They were admitted in early 2021 with urosepsis and managed with broad spectrum antibiotics. HIV-1 was diagnosed 20 years previously and the patient had a history of poor adherence to ART. Four weeks into the admission, a CNTS was positive for SARS-CoV-2 RNA by RT-PCR. The patient required a short ITU admission for high flow oxygen. All subsequent CNTS following this have been positive for SARS-CoV-2 RNA by RT-PCR.

Patient C is in their 30s and was also admitted in early 2021 with a history of cough, fever and odynophagia. HIV-1 had been diagnosed over 10 years previously and adherence to ART had been intermittent since diagnosis. SARS-CoV-2 RT-PCR on a CNTS was positive on admission. The patient was swabbed and tested weekly for SARS CoV-2 and although asymptomatic since admission, all CNTS to date are SARS-CoV-2 positive.

## RESULTS

Figure 1 shows HIV-1 viral load, CD4 cell count, ART regimen and anti-S IgG antibody titre and amino acid mutations in the receptor binding domain (RBD) of S-protein that have previously been observed in variants of concern (VOC), that arose in these patients over time and were detected in subsequent samples. Amino acid changes are either seen at the consensus level or are part of a mixed viral population.

**Figure 1.**
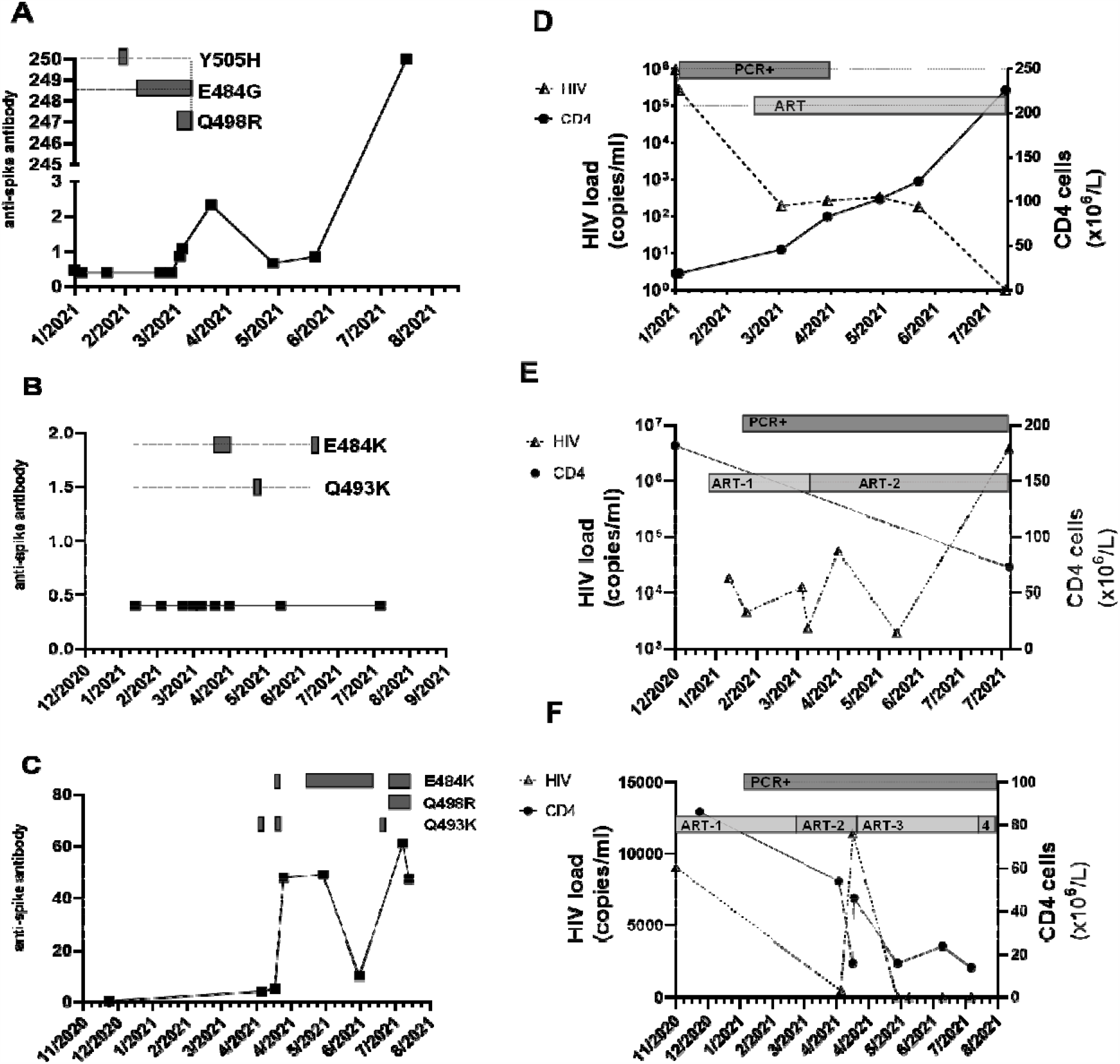
Serial CNTS samples from (a; d) patient A; (b; e) patient Band (c; f) patient C. Anti-spike IgG was measured using commercial assay (Roche Elecys anti-SARS-COV-2 anti –Spike antibody), units of measure are arbitrary, cut-off for positive result indicated by dashed horizontal line. Amino acid changes in the receptor binding domain (RBD) of the spike protein at locations previously associated with variants of concern (VOC) that arose over the same time period are displayed as boxes along the dotted horizontal line. Amino acid changes are either seen at the consensus level or are part of a mixed viral population. In addition, the alpha lineage mutation N501Y was present in all samples. HIV-1 viral load was measured using the Roche Cobas 8800, and CD4 cell count were measured and the SARS-COV-2 RT-PCR positivity and ART regimen over the same time is also displayed. (d) ART: Trimeq (DTG/ABC/3TC)+DTG; (e) ART-1: Bictarvy (BTG/TAF/ETC), ART-2: TAF/ETC/DRV/c; (f) ART-1: TDF/ETC/DRV/r; ART -2 : TAF/ETC/DRV/c; ART-3: TAF/ETC/DTG; ART-4: TDF/ETC/DTG

Clinical and laboratory data (including CD4 counts and HIV viral load) for patients A, B and C was collected during their inpatient stay (supplementary table S1).

SARS-CoV-2 genome sequencing was performed on all positive samples. In all cases, sequencing confirmed B.1.1.7 (alpha) lineage. The amino acid changes in relation to Wuhan Hu-1 SARS-CoV-2 are presented in supplementary table S2.

We analysed anti-S IgG using serum and ancestral strain virus neutralisation for Patient C. All serum samples tested were unable to neutralise Wuhan Hu-1 SARS-CoV-2 (IC50< 50, supplementary figure S3. A) despite detection of anti-S IgG (>0.8, Supplementary Figure S3.B).

## DISCUSSION

Three patients, all with advanced HIV-1 infection, CD4 lymphopenia and chronic SARS-CoV-2 infection are described.

Persistent detection of SARS-CoV-2 RNA is common and does not always correspond with productive infection. All three patients had sufficient viral RNA detected in CNTS sampled over several months of follow up for successful genome sequencing. This typically corresponds to a Ct value less than 30 in the majority of RT-PCR assays, and although these assays are frequently not quantitative, this normally equates to a moderate or high viral load (7). In all three patients we observed the generation of novel mutations over time, some of which persisted (Fig. 1). This persistence is presumably due to selection pressure, and therefore is evidence of active viral replication. In addition, the virus was successfully cultured from one of patient C’s samples in September 2021. These findings provide further evidence that in the setting of immunosuppression, individuals can have chronic SARS-CoV-2 infection providing an opportunity for the generation of novel variants.

Cellular studies suggest that the mutations we have observed in the recovered SARS-CoV-2 genomes from these patients will enhance transmissibility and may be associated with antibody escape (8).

All patients were infected with the B.1.17 (alpha) lineage that contains N501Y, a mutation associated with an approximately 10 fold higher binding affinity of the RBD for ACE2 (9). It is notable that all three patients developed a mutation at position E484 which is located in the receptor binding motif (Fig 1. A, B and C). The emergence of these mutations in genomes from B.1.1.7 lineage has previously been described (10). Substitutions at E484, including E484K and E484G, enhance affinity of binding to ACE2(11). These mutations have been shown to significantly affect neutralisation with therapeutic antibodies and are a major concern for vaccine escape (12). N501Y and E484K are together present in variant B.1.351 where a significant decrease in neutralising antibody activity from the sera of vaccinated individuals has been observed (13). *In vitro* studies predict the combination of Q498R with N501Y significantly enhance human ACE2 binding (14). The summative deleterious effects of mutations N501Y, Q498R and E484K/G observed in patients A and C (Fig 1. A and C) are also predicted *in vitro* to increase ACE2 binding by 50-fold, compared with WT which may translate into increased transmissibility (14). This suggests significant potential for immune evasion however, this would need to be characterised further with *in vitro* and *in vivo* studies. Q493K (Fig 1. B and C) located in the RBD has previously been detected in immunocompromised hosts and has been shown to lead to resistance to nAb (15). The effect of Y505H (Fig 1. A) is unknown but it is located in the receptor binding motif (16). Interestingly, Q493K, Q498R and Y505H are mutations identified in the omicron SARS-CoV-2 variant.

There was no neutralisation against WT-Wuhan SARS-CoV-2 detected from sera at different time points from patient C (supplementary Fig. S3). It is extrapolated that there will also be poor neutralisation of SARS-CoV-2 B.1.1.7 lineage. The lack of neutralising ability in patient C despite detectable anti-S antibodies via commercial methods emphasizes the importance of a functional assessment of the immune response in immunocompromised individuals.

It is not possible to determine whether the lack of neutralising antibody, CD4 lymphopenia or a selection advantage due to enhanced receptor binding led to the mutations observed in these patients. However it is possible that ineffective neutralising antibody responses are a consequences of CD4 cell lymphopenia as neutralising antibody responses are largely T cell dependent (17, 18). Also CD4 T cells have been shown to be associated with recovery from SARS-CoV-2 infections (18). SARS-CoV-2 specific CD4 T cells have also been associated with accelerated viral clearance (19). The observed chronic SARS-CoV-2 infections in our patients support this. However absent/delayed CD4 cell responses have also been associated with severe or fatal disease where our patients remained largely asymptomatic (19). These findings may indicate the role of other arms of the adaptive immune response.

## CONCLUSIONS

These three immunocompromised patients are further examples of chronic SARS-CoV-2 infection. CD4 T-cells are central to both neutralising antibody and cytotoxic T-cell activity in acute infection. Indeed our patients, all with CD4 lymphopenia, developed a chronic infection. This supports a critical role for CD4 T-cells in the clearance of SARS-CoV-2 infection. Our observations highlight the need for further research to understand the consequences of specific immune deficits in control of SARS-CoV-2 and other (viral) infections.

Early identification and detection of patients with chronic SARS-CoV-2 infection is essential to prevent transmission of variants with potential for vaccine escape. Our patients highlight a novel use of viral genome sequencing data to identify replication competent virus which subsequently enabled appropriate Infection Prevention and Control measures to be deployed. This suggests access to rapid viral sequencing in clinical diagnostic laboratories is essential in the management of this and future epidemics.

If or how these patients with chronic infection influence the emergence of new variants is unclear. These chronically infected patients pose a significant risk in resource poor settings: identification of them is challenging due to reduced access to healthcare, RT-PCR and NGS, the majority of the population remain unvaccinated and in sub-Saharan Africa there is a higher burden of people living with HIV (around 30% who are not receiving appropriate ART) (20). This underlines the moral imperative to ensure equity of access to COVID vaccines across the World.

## Data Availability

All data produced in the present study which is not included in the manuscript and supplementary material are available upon reasonable request to the authors

http://cov-glue.cvr.gla.ac.uk/#/home

## ACKNOWLEDGEMENTS

Thanks to the Rare and Imported Pathogens Laboratory, Salisbury, UK for virus culture.

