## Supplementary material for "Generation of novel SARS-CoV-2 variants on B.1.1.7 lineage in three patients with advanced HIV disease"

**MATERIALS AND METHODS**

SARS-CoV-2 RNA testing was performed using two platforms. The Hologic® Aptima® SARS-CoV-2 assay is a thermal mediated amplification (TMA) method with two targets in the ORF1AB gene of SARS-CoV-2. This reports detection (or no detection) via reactive light units (RLU). With this method it is not possible to extrapolate the quantity of viral RNA in the sample. The second platform used for detection of SARS-CoV-2 RNA from this patient’s samples was the Roche Cobas® 8800 which is a standard automated real time RT-PCR with two gene targets, the ORF1A and E gene.

Whole Genome Sequencing (WGS) of the viral genome was performed *in house* on stored aliquots of the samples collected from all three patients. Samples with a RT-PCR cycle threshold (CT) value < 32 in the Roche Cobas® 8800 assay contain sufficient viral RNA for sequencing. Sequencing was also attempted from samples with an RLU > 1,000 in the Hologic® Aptima® SARS-CoV-2 assay, however these will include a proportion of sample with insufficient viral RNA for sequencing. Briefly, samples were sequenced with a multiplex PCR-based approach according to the modified ARTIC protocol with version 3 primer set. Amplicon libraries (Nextera v2) were sequenced using Illumina MiSeq and Illumina MiniSeq. Genomes were assembled with reference based assembly (<https://artic.network/ncov-2019/ncov2019-bioinformatics-sop.html>) with 10x minimum coverage cut-off for any region of the genome and 50% cut-off for defining single nucleotide polymorphisms (SNPs). The lineage was determined by using Pangolin lineage assignment tool^[1]^. The generated FASTA files were uploaded to the CoV-GLUE (<http://cov-glue.cvr.gla.ac.uk/#/home>) web platform for further analysis.

All HIV-1 viral loads were performed on the Roche COBAS 6800 platform. This is a real time PCR assay targeting two unique regions of the HIV-1 genome, gag and LTR. The LOD of this assay is 50 copies per mL in our laboratory settings.

The anti-spike antibody responses were evaluated by testing stored sera from each of the patients. We used the Roche Elecsys anti-SARS-COV-2 anti –Spike antibody; a double antigen sandwich assay targeting the receptor binding domain of the S antigen. The cut off for detection of this assay is 0.8 U/mL.

SARS-CoV-2 microneutralisation assay was performed as described previously (Reynolds et al. 2020 and Reynolds et al 2021). Vero E6 cells were seeded in 96-well plates 24h prior to infection. Duplicate titrations of heat-inactivated patient sera were incubated with 3x10^4^ FFU SARS-CoV-2 virus (TCID100) at 37°C for 1h. Serum/virus preparations were added to cells and incubated for 72h. Surviving cells were fixed in 3.7% (vol/vol) formaldehyde and stained with 0.1% (wt/vol) crystal violet solution. Crystal violet stain was resolubilised in 1% (wt/vol) sodium dodecyl sulphate solution. Absorbance readings were taken at 570nm using a CLARIOStar Plate Reader (BMG Labtech). Negative controls of pooled pre-pandemic sera (collected prior to 2019), and pooled serum from neutralisation positive SARS-CoV-2 convalescent individuals were spaced across the plates. Absorbance for each well was standardised against technical positive (virus control) and negative (cells only) controls on each plate to determine percentage neutralisation values. IC50s were determined from neutralisation curves. All authentic SARS-CoV-2 propagation and microneutralisation assays were performed in a containment level 3 facility.

1. *Toole, A et al. Assignment of epidemiological lineages in an emerging pandemic using the pangolin tool. Virus Evolution 2021.*
