## Supplementary tables and figure for "Generation of novel SARS-CoV-2 variants on B.1.1.7 lineage in three patients with advanced HIV disease"

| **Patient** | **Sample date** | **HIV VL (copies/ml)** | **CD4 (x10^6^/L)** | **SARS-CoV-2 RT-PCR** | **Anti-Spike antibody** | **Anti-Nucleocapsid antibody** |
| --- | --- | --- | --- | --- | --- | --- |
| **A** | January 2021 | 975, 000 | 19 | Positive | ND | ND |
|  | February 2021 | 269, 000 | 19 | Positive | ND | ND |
|  | March 2021 |  |  |  | ND | ND |
|  | March 2021 | 194 | 46 | Positive | D | D |
|  | April 2021 | 268 | 83 | Positive | D | D |
|  | May 2021 | 333 | 103 | Negative | ND | D |
|  | June 2021 | 186 | 123 | Negative | D | D |
|  | July 2021 | 0 | 226 | Negative | D | D |
| **B** | December 2020 |  | 182 | Negative |  |  |
|  | January 2021 | 18, 700 |  | Negative |  |  |
|  | February 2021 | 4, 550 |  | Positive | ND | ND |
|  | March 2021 | 12, 600 |  | Positve | ND | ND |
|  | March 2021 | 2, 330 |  | Positve | ND | ND |
|  | April 2021 | 55, 800 |  | Positve | ND | ND |
|  | May 2021 | 1920 |  | Positve | ND | ND |
|  | August 2021 | 3, 790, 000 | 73 | Positive | ND | ND |
| **C** | December 2020 |  | 86 | Positive | ND | ND |
|  | April 2021 | <100 | 54 | Positive | D | ND |
|  | April 2021 | 449 | 16 | Positive | D | ND |
|  | May 2021 | 0 | 16 | Positive | D | ND |
|  | June 2021 | 0 |  | Positive | D | ND |
|  | July 2021 | 0 | 14 | Positive | D | ND |
|  | August 2021 |  |  |  | D | ND |

**Supplementary Table S1.** HIV viral load, CD4 count, SARS-CoV-2 RT-PCR and anti-S and anti-N antibody results for patients A, B and C

D = detected	ND = not detected

| Patient | Sample date | COG ID | GISAID ID | ENA ID | SNPs Total | SNPs in S | Identified spike amino acid substitutions | | | | | | | | | | | | B.1.17 defining mutations | | | | |
| --- | --- | --- | --- | --- | --- | --- | --- | --- | --- | --- | --- | --- | --- | --- | --- | --- | --- | --- | --- | --- | --- | --- | --- |
|  |  |  |  |  |  |  | F79 | T95* | G142* | D215† | S254 | L455 | E484 | F486 | Q493* | Q498* | N501Y | A570D | D614G | P681H | T716I | S982A | D1118H |
| A | February 2021 | LOND-128D956 | EPI_ISL_1730308 | ERR5776569 | 37 | 7 |  |  |  |  |  |  |  |  |  |  | x | x | x | x | x | x | x |
|  | February 2021 | LOND-128D983 | EPI_ISL_1730311 | ERR5777434 | 38 | 8 |  |  |  |  |  |  |  |  |  |  | x | x | x | x | x | x | x |
|  | March 2021 | LOND-128D9A1 | EPI_ISL_1730313 | ERR5777502 | 45 | 9 |  |  |  |  |  |  |  |  |  |  | x | x | x | x | x | x | x |
|  | March 2021 | LOND-128D9B0 | EPI_ISL_1730314 | ERR5777044 | 41 | 9 |  |  |  |  |  |  | EG |  |  |  | x | x | x | x | x | x | x |
|  | March 2021 | LOND-128D455 | EPI_ISL_1636965 | ERR5747961 | 37 | 10 |  |  |  |  |  |  | G |  |  | R | x | x | x | x | x | x | x |
|  | April 2021 | LOND-128DA35 | High QC Failed | ERR5776980 | 29 | 7 |  |  |  |  |  |  | G |  |  | R | x | x | x | x | x |  |  |
| B | February 2021 | LOND-128D965 | EPI_ISL_1730309 | ERR5777875 | 31 | 7 |  |  |  |  |  |  |  |  |  |  | x | x | x | x | x | x | x |
|  | February 2021 | LOND-128D974 | EPI_ISL_1730310 | ERR5776147 | 36 | 9 |  |  |  |  |  |  |  |  |  |  | x | x | x | x | x | x | x |
|  | March 2021 | LOND-128D992 | EPI_ISL_1730312 | ERR5775954 | 32 | 9 |  |  | GV |  | SF |  |  |  |  |  | x | x | x | x | x | x | x |
|  | March 2021 | LOND-128D9CF | EPI_ISL_1730315 | ERR5777489 | 31 | 8 |  |  |  |  | SF |  |  |  |  |  | x | x | x | x | x | x | x |
|  | March 2021 | LOND-128D9DE | EPI_ISL_1730316 | ERR5777971 | 35 | 10 |  | TI | GV |  | SF |  |  |  |  |  | x | x | x | x | x | x | x |
|  | April 2021 | LOND-128DA17 | EPI_ISL_1730319 | ERR5777283 | 35 | 10 | LF |  |  |  | SF |  | KE^§^ |  |  |  | x | x | x | x | x | x | x |
|  | April 2021 | LOND-128DA26 | EPI_ISL_1730320 | ERR5776220 | 35 | 10 | LF |  |  |  | SF |  | KE^§^ |  |  |  | x | x | x | x | x | x | x |
|  | April 2021 | LOND-128DF45 | EPI_ISL_2126006 | ERR5941392 | 35 | 9 |  |  |  |  | SF |  |  |  |  |  | x | x | x | x | x | x | x |
|  | May 2021 | LOND-128E81C | EPI_ISL_2355373 | ERR5989922 | 39 | 11 | LF |  |  |  | SF |  |  |  | K |  | x | x | x | x | x | x | x |
|  | June 2021 | LOND-128E973 | EPI_ISL_2517310 | ERR6063636 | 44 | 10 | LF |  |  |  | SF |  | KE^§^ |  |  |  | x | x | x | x | x | x | x |

| Patient | Sample date | COG ID | GISAID ID | ENA ID | SNPs Total | SNPs in S | Identified spike amino acid substitutions | | | | | | | | | | | | B.1.17 defining mutations | | | | |
| --- | --- | --- | --- | --- | --- | --- | --- | --- | --- | --- | --- | --- | --- | --- | --- | --- | --- | --- | --- | --- | --- | --- | --- |
|  |  |  |  |  |  |  | F79 | T95* | G142* | D215† | S254 | L455 | E484 | F486 | Q493* | Q498* | N501Y | A570D | D614G | P681H | T716I | S982A | D1118H |
| C | January 2021 | LOND-128D947 | EPI_ISL_1730307 | ERR5777630 | 30 | 7 |  |  |  |  |  |  |  |  |  |  | x | x | x | x | x | x | x |
|  | March 2021 | LOND-128A92C | EPI_ISL_1474822 | ERR5666834 | 33 | 8 |  |  |  |  |  |  |  |  |  |  | x | x | x | x | x | x | x |
|  | March 2021 | LOND-128D9FC | EPI_ISL_1730318 | ERR5776269 | 37 | 11 |  |  |  |  | SF | LF |  |  | K |  | x | x | x | x | x | x | x |
|  | April 2021 | LOND-128DA08 | High QC Failed | ERR5776785 | 33 | 5 |  |  |  |  |  |  |  |  |  |  |  |  |  | x | x | x |  |
|  | April 2021 | LOND-128F9FA | EPI_ISL_3771691 | ERR6633239 | 39 | 9 |  |  |  |  |  |  |  |  |  |  | x | x | x | x | x | x | x |
|  | April 2021 | LOND-128DD1E | EPI_ISL_1975060 | ERR5938419 | 54 | 12 |  |  |  |  |  | LF | KE^§^ | IF | KQ |  | x | x | x | x | x | x | x |
|  | May 2021 | LOND-128DF90 | EPI_ISL_2126013 | ERR5938492 | 37 | 9 |  |  |  |  |  |  |  |  |  |  | x | x | x | x | x | x | x |
|  | May 2021 | LOND-128E560 | EPI_ISL_2238913 | ERR5971254 | 34 | 8 |  |  |  |  |  |  |  |  |  |  | x | x | x | x | x | x | x |
|  | May 2021 | LOND-128FA06 | High QC Failed | ERR6627346 | 34 | 10 |  |  |  |  |  |  |  |  |  |  | x | x | x | x | x | x | x |
|  | May 2021 | LOND-128FA15 | EPI_ISL_3771692 | ERR6621356 | 42 | 12 |  | TI |  |  | SF |  | KE^§^ |  |  |  | x | x | x | x | x | x | x |
|  | June 2021 | LOND-128FA24 | EPI_ISL_3771693 | ERR6629896 | 46 | 14 |  | TI |  | DG | SF | LF | K^§^ | FI |  |  | x | x | x | x | x | x | x |
|  | June 2021 | LOND-128FA33 | EPI_ISL_3771694 | ERR6625218 | 46 | 14 |  | TI |  |  | SF | LF | K^§^ | FI |  |  | x | x | x | x | x | x | x |
|  | July 2021 | LOND-128F408 | EPI_ISL_3420518 | ERR6541328 | 51 | 12 |  |  |  |  |  |  | K^§^ | I |  |  | x | D Y | x | x | x | x | x |
|  | July 2021 | LOND-128F417 | EPI_ISL_3420519 | ERR6631229 | 49 | 15 |  | TI | GV | DY | SF | LF | K^§^ | FI |  | R | x | x | x | x | x | x | x |
|  | August 2021 | LOND-128F426 | EPI_ISL_3420520 | ERR6541316 | 50 | 13 |  |  | GV |  | SF |  | K^§^ | IF |  | QR | x | D Y | x | x | x | x | x |
|  | August 2021 | LOND-128FBB8 | EPI_ISL_3771717 | ERR6622262 | 43 | 14 |  | I | GV |  | F | F | K^§^ |  |  | R | x | x | x | x | x | x | x |
|  | August 2021 | LOND-1290237 | EPI_ISL_4113884 | ERR6765764 | 48 | 15 |  | TI | GV |  |  |  | K^§^ | I |  | QR | x | x | x | x | x | x | x |
|  | September 2021 | LOND-1290729 | EPI_ISL_4769357 | ERR7002956 | 51 | 16 |  | TI | GV | DG | SF |  | K^§^ | I |  | R | x | x | x | x | x | x | x |
|  | September 2021 | LOND-129099C | EPI_ISL_5039388 | ERR7049253 | 52 | 14 |  |  | V | DY |  |  | K^§^ | I |  | R | x | x | x | x | x | x | x |
|  | September 2021 | LOND-1290AE4 | EPI_ISL_5265186 | ERR7143334 | 43 | 14 |  | TI | V |  | F | LF | K^§^ |  |  | R | x | x | x | x | x | x | x |

**Supplementary Table S2.** Amino acid substitutions and SNPs identified via NGS of respiratory samples from patients A, B and C. Where two amino acids are recorded this represents a mixed viral population containing both amino acids at this position. * = amino acid substitutions in Omicron variant (B.1.1.529) † = Beta variant (B.1.351) defining amino acid substitution § = Mu (B.1.621.1) and Gamma (P.1) defining amino acid substitutions x = detected

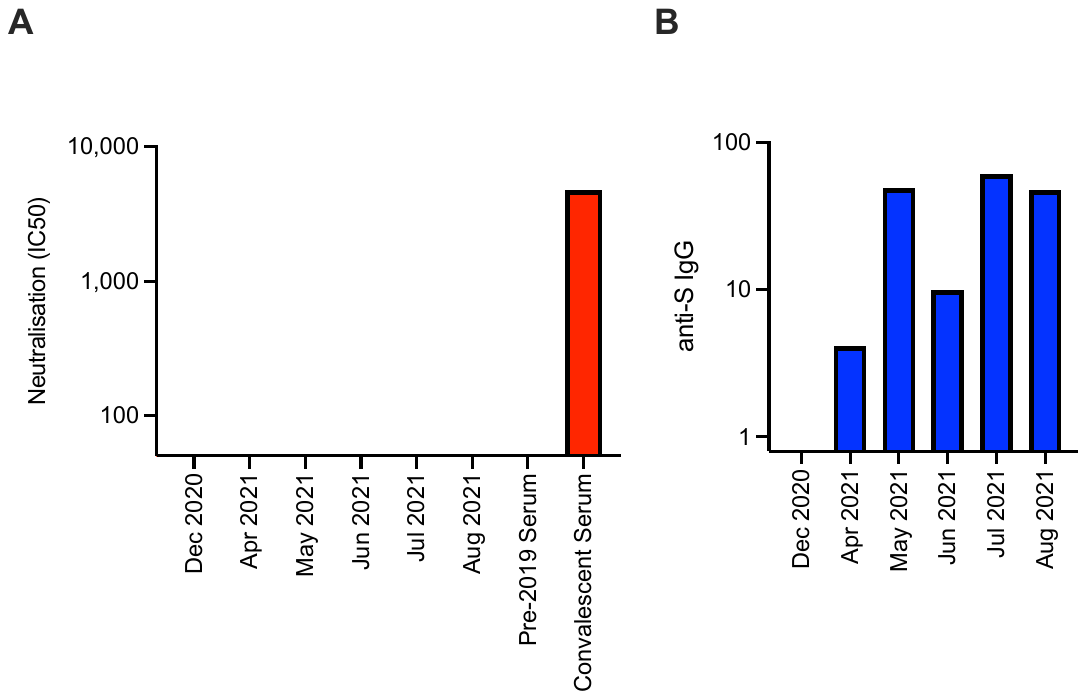

**Supplementary Figure S3. A**, Neutralisation titres (IC50) for serum samples from Patient C (dates indicated). Pre-pandemic (pre-2019) human serum and pooled convalescent serum were included as controls. **B**, anti-S IgG titres in serum samples from Patient C (dates indicated).
